# Dietary inflammatory index scores and cognitive aging: results from the Framingham heart Study offspring cohort

**DOI:** 10.1101/2024.11.27.24318084

**Authors:** Debora Melo van Lent, Paul F. Jacques, Sokratis M. Charisis, Hannah Gokingco Mesa, Claudia Satizabal, Changzheng Yuan, Ramachandran S. Vasan, Sudha Seshadri, Alexa Beiser, Jayandra J. Himali, Mini E. Jacob

## Abstract

**Background:** Nutritional factors can abet or protect against systemic chronic inflammation, which plays an important role in the development and progression of dementia. We evaluated whether higher (i.e. pro-inflammatory) Dietary Inflammatory Index (DII) scores were associated with cognitive decline in the community-based Offspring Framingham Heart Study (FHS).

**Method:** 1,614 older adults (mean age 61 years [standard deviation (SD)], 9;55% women]) completed validated 126-item Food Frequency Questionnaires (FFQ), administered at FHS examination cycle 7 (1998-2001) and examination cycle 5 (1991-1995) and/or 6 (1995-1998). We created a DII score (based on the published method by Shivappa et al. 2014) for each available exam; a cumulative DII score was calculated by averaging exam-specific scores across the two or three exams. Cognitive testing was completed at call back sessions following examination cycles 7 and 8 (2005-2008). Exam 7 was considered as study baseline and participants were followed over a mean time of seven years (SD 1). We excluded participants with prevalent dementia at baseline and those with no cognitive testing data at either/or exams 7 and 8. We examined associations between the cumulative DII score and cognitive test scores over time using annualized change adjusting for age, sex, education (model 1) and additionally for exam 7 measures of body mass index, total energy intake, total cholesterol: high-density lipoprotein ratio, smoking and anti-cholesterol medication (model 2).

**Results:** Higher DII scores were significantly associated with (less) decline in performance on Similarities (verbal comprehension/reasoning) and Global cognition, following adjustments for model 1 covariates (Model 1:β and SE, 0.017, 0.008,p=0.03; 0.004,0.002,p=0.02, respectively). The effect sizes remained similar after additional adjustment for Model 2 covariates (0.018, 0.009,p=0.06; 0.004, 0.002,p=0.03, respectively). Additionally, we found that higher DII scores associated with accelerated decline in performance on Trail Making B-A (processing speed and executive function) (Model 2: −0.010, 0.004,p=0.03). We observed no relationships between higher DII scores and other neuropsychological tests. Further, stratified analyses revealed a linear relationship between higher DII scores and (less) decline in performance on Hooper visual organization among men, but not women (Model 2: 0.022, 0.010,p=0.02; −0.011, 0.009,p=0.23, respectively).

**Conclusion:** Higher DII scores were associated with (less) cognitive decline. We take our unexpected findings with caution as we previous have seen a relationship between higher DII scores and increased risk for dementia. To date, such studies have been very limited, most studies that found a relationship were cross-sectional and have used less sensitive testing. Future longitudinal studies with sensitive neuropsychological test measures are encouraged to elucidate whether a longitudinal relationship between higher DII scores and age-related cognitive decline exists.

## Introduction

Aging is a key non-modifiable risk factor for most non-communicable diseases and reduces our ability to fully function in older age. Cognition is one of those abilities that naturally declines with age^1^. In addition, ageing-associated cognitive decline is affected by organ systems and processes, for example, the immune system^2^. Indeed, aging-induced (low grade) systemic inflammation – inflammageing – has been associated with accelerated cognitive decline^3^ ^4^. Inflammageing is hypothesized to be caused by processes including cellular senescence and dysregulation of innate immunity, and characterized by elevated blood concentrations of pro-inflammatory cytokines such as interleukin-6 (IL-6) and tumor necrosis factor-alpha (TNF-α) in older age^5^. Consequently, inflammageing contributes to ageing related outcomes such as cognitive decline and brain aging^4^ ^6^.

Modifiable disease risk factors, such as diet, may combat inflammageing and consequently slow cognitive decline. Evidence from clinical trials has shown that diet may play a key role in reducing systemic inflammation. For example, the Prevención con Dieta Mediterránea (PREDIMED) randomized control trial (RCT) reported that long term adherence to the Mediterranean diet - a diet rich in anti-inflammatory foods and food groups - significantly lowered concentrations of important inflammatory markers such as: IL-6, TNF-α and high-sensitivity C-reactive protein (CRP) ^7^. In addition, long term adherence to the (energy restricted) Mediterranean diet and the Mediterranean diet in combination with extra-virgin olive oil were associated with better performance on language and memory tests^8^ ^9^.

A limitation, however, of predefined dietary patterns such as the Mediterranean diet, is that it is specific to the European Mediterranean region. To extrapolate the diet to other populations is challenging, as foods included in the diet may not be available or not part of the country specific dietary habits. A more effective way to optimize dietary guidelines across the globe is to assess inflammatory content of diet through a dietary index primarily based on nutrients (e.g. vitamin D) and global available foods (e.g. black tea). Results from such nutrient and global food indices can be translated into foods and food groups available in regional dietary guidelines.

The Dietary Inflammatory Index (DII), is an index that has been specifically designed to capture the inflammatory potential of diet across the globe^10^. The index has been associated with a wide number of health outcomes, including incident dementia^11^ ^12^.

In our Framingham Heart Study (FHS) Offspring cohort, we found that higher DII scores were associated with increased incidence of all-cause and Alzheimer’s Disease (AD) dementia^11^, and with global brain volume measures, such as smaller total brain volume and total gray matter volume^13^. As we have seen relationships with dementia and brain aging, we are interested in the relationship between DII and cognitive aging. The current evidence of the relationship between the DII and cognitive aging is primarily based on studies that used the Mini-Mental State Examination (MMSE) or the Montreal Cognitive Assessment (MOCA) ^14–17^. A limited number of studies used sensitive neuropsychological measures^18–23^, but exposure and outcomes were 13 years apart^18^, had a cross-sectional study design^19–21^ ^23^, were carried out in women only^22^ and/or was carried out in small study samples^19^ ^23^. A large cohort, with longer follow-up time, usual dietary intake measured over consecutive visits and sensitive neuropsychological measures would provide substantial evidence for the role of diet on cognitive aging. Therefore, we examined the relationship between the DII and cognitive decline in a large established cohort with repeated diet measures and adequate follow-up time. We studied the association between the DII and cognitive performance over a period up to 11 years follow-up in 1614 participants of the community-based Framingham Heart Study (FHS) Offspring cohort. A positive association, if detected, would imply that dietary intervention focusing on anti-inflammatory content of diet could be part of public health preventive strategies to combat ageing-associated cognitive decline across the globe.

## Methods

The FHS is an ongoing community-based study of several cohorts from the town of Framingham, Massachusetts, USA (https://www.framinghamheartstudy.org/). The Original cohort was established in 1948 with the aim to identify factors that contribute to the development of cardiovascular disease^24^. In 1971, the Offspring cohort was established, including children of the Original cohort and their spouses^25^. The Offspring cohort enrolled 5124 participants who have been studied over ten examination cycles, approximately once every 4 years. All participants provided written informed consent. The study protocol was approved by the institutional review board at Boston University Medical Center.

For the present study, we assessed self-reported dietary intake from participants of the Offspring cohort using a food frequency questionnaire (FFQ) administered at examination cycles 5 (1991- 1995), 6 (1995-1998), and 7 (1998– 2001). A flow chart of sample selection is shown in **Figure 1**. To be included in the present investigation, participants were required to have completed the FFQ at examination cycle 7 and at least one other time point (examination cycles 5 or 6). Participants were excluded if they had no dietary intake data available or an abnormal estimated total energy intake (<600 - >3999 kcal for women or <600 - >4199 kcal for men), and/or >13 missing FFQ items (n=643)^26^. In addition, we excluded participants with no cognitive test data (exam 7: 1998-2001 (n=349)) at baseline, participants with no cognitive test data at follow-up (exam 8: 2005-2008 (n= 931)). or prevalent dementia (n=2).

**Figure 1.**
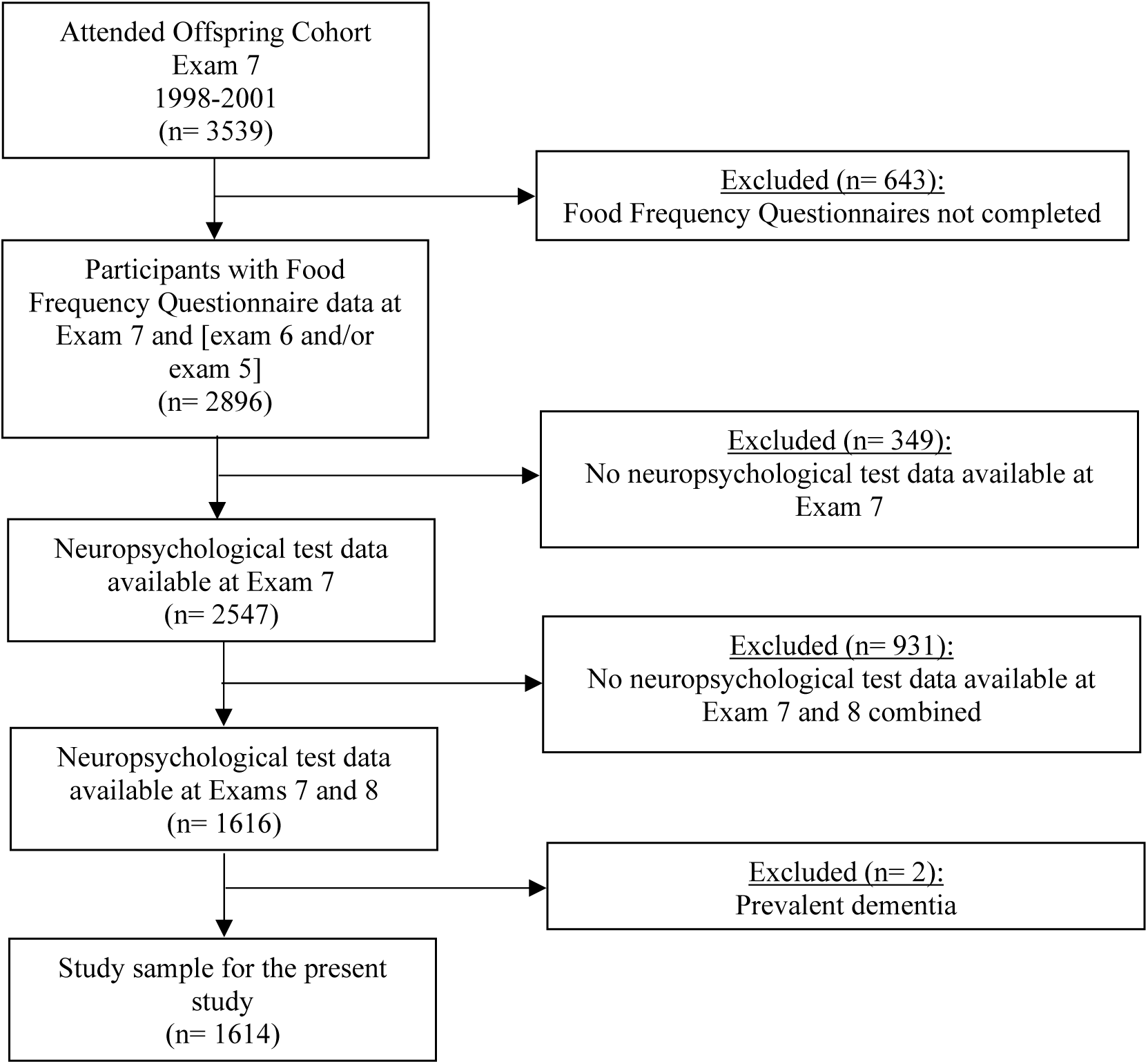
Flow chart of the participants included in the study.

### Dietary inflammatory index scores

The DII index for the present study was calculated using the validated 126-item Harvard semi-quantitative FFQ^27^ ^28^. The FFQ assesses dietary intake over the past year. Participants were asked how often they consumed food items (i.e., from never or <1 per month to >6 per day), the type of food item and whether food items were homemade or ready-made ^29^. A commonly used portion size was given for each food item. In addition, the FFQ included questions on most frequently eaten breakfast cereal, types of fats and oils, and frequency of consumption of fried foods. Intakes of food components (i.e. nutrients, food items or food groups) were computed by multiplying the frequency of consumption of each food item by the nutrient content of the specified portions^29^.

The DII index for the present study consists of 36 dietary components including anti-inflammatory nutrients, pro-inflammatory nutrients, whole foods and caffeine (5). The dietary components are categorized as (1) *Anti-inflammatory*: alcohol, beta-carotene, caffeine, dietary fiber, folic acid, magnesium, thiamin, riboflavin, niacin, zinc, monounsaturated fat, polyunsaturated fat, omega-3 fat, omega-6 fat, selenium, vitamins B6, A, C, D, E, flavan-3-ol, flavones, flavonols, flavonones, anthocyanidins, green/black tea, pepper, and garlic; and (2) *pro-inflammatory*: vitamins B12, iron, carbohydrates, cholesterol, total energy intake, protein, saturated fat, and total fat. Nine components of the Shivappa et al. 2014 DII (i.e. turmeric, thyme/oregano, rosemary, eugenol, ginger, onion, transfat, isoflavones, and saffron) were not available in our FFQ. The algorithm for the DII computation has been described elsewhere **(eTable 1 in the supplement)**^10^.

At FHS we had the opportunity to assess dietary intake over a decade. As data from a FFQ is prone to recall bias, and to account for reverse causality, we averaged the DII scores across examination cycle 7 (1998-2001) and at least one of the prior examination cycles: 5 (1991-1995) and 6 (1995-1998).

### Cognitive decline

Cognitive decline (assessed as annualized change between exam 7 and 8) was measured using a battery of validated neuropsychological (NP) tests. Our battery included Visual Reproductions Delayed Recall (visual memory) and Logical Memory Delayed Recall (meaningful/verbal memory) from the Wechsler Memory Scale, Similarities (verbal comprehension/reasoning) from the Wechsler Adult Intelligence Scale, Trail Making Test A (processing speed) and Trail Making Test B minus A (processing speed and executive function), and the Hooper Visual Organization Test (closure/visual integration and mental rotation) ^30^. We also used principal component analysis (i.e. forcing a single component solution. Task scores were standardized and subsequently being summed together according to their weighting to the overall cognitive factor^31^) to investigate a composite measure of cognitive function reflecting general cognitive ability. The score combines weighted loadings for Trail-Making Test Part B, Logical Memory, Visual Reproductions, and Similarities. Higher scores across all cognitive endpoints indicate superior performance, except Trail-Making Test, whereby higher scores indicate slower task completion. For the Trail Making Test scores directionality was reversed such that higher scores on all tests reflected less decline in performance (prospectively).

### Statistical analysis

SAS Software 9.4 (SAS Institute, Cary, NC, USA) was used to perform separate multivariable linear regressions to examine the associations between the DII and outcomes.

The main results are presented as adjusted beta coefficients accompanied by standard errors. The beta estimates represent change in annualized change of each respective NP test per one unit increase in the DII score, negative beta indicate more decline. Annualized change was calculated by subtracting NP tests at exam 7 from NP tests at exam 8 and dividing by time in years (i.e. time between NP exam dates divided by 365.25). The DII was analyzed as both a continuous variable and categorical variable (i.e. quartiles) with quartile 1 as the reference category. *P*-values <0.05 and <0.10 was considered statistically significant for our primary analysis and for our tests of interactions, respectively. Missing data were excluded from analysis. Covariates were selected based on published literature **(eTable 2 in the Supplement)**. Model 1 was adjusted for age, sex and education. Model 2 was additionally adjusted for lifestyle factors (body mass index (BMI), total energy intake, and smoking status), cardiometabolic factors (total to high-density lipoprotein (HDL) cholesterol ratio and the use of anti-cholesterol medication). For our secondary analyses, we tested for interactions between the DII and sex separately using model 2.

## Results

### Cohort demographics

Table 1 details the sample characteristics. Participants were on average 61 years old at baseline (standard deviation 9) and 55% were women. The mean DII score was −0.24 ± 1.66, indicating that this sample’s diets were on average anti-inflammatory relative to the global mean. Follow up time between exam 7 and 8 ranged between 1 – 11 years (median time 7 years).

**Table 1.** Baseline characteristics of study sample (n=1614)

| Characteristic<sup>1</sup> | Overall study sample |
| --- | --- |
|  | N=1614 |
| Age, years | 61 ± 9 |
| Women, n (%) | 894 (55) |
| Education, n (%) |  |
| No high school degree | 48 (3) |
| High school degree | 413 (26) |
| Some years of college | 483 (30) |
| College degree | 670 (42) |
| Body Mass Index (kg/m<sup>2</sup>) | 27.9 ± 5.3 |
| Current smokers | 157 (10) |
| Total caloric intake | 1859 ± 593 |
| Total to HDL Cholesterol ratio | 4.0 ± 1.3 |
| Usage of anti-cholesterol medication, n (%) | 306 (19) |
| Dietary Inflammatory Index score | -0.25 ± 1.66 |
| Neuropsychology Measures |  |
| Visual reproduction delayed, n correct | 8.3 ± 3.3 |
| Logical memory delayed, n correct | 10.8 ± 3.5 |
| Trail Making A, minutes, Median [Q1, Q3] | 0.5 [0.4, 0.6] |
| Trail Making B-A, minutes, Median [Q1, Q3] | 0.6 [0.4, 0.9] |
| Hooper Visual Organization Test, total | 26.0 [24.0, 27.0] |
| Correct, Median [Q1, Q3] |  |
| Similarities | 17.1 ± 3.4 |
| Global cognition | 0.14 ± 0.91 |
| Follow-up time, years | 7 ± 1 |

Mean ± SD reported, unless specified otherwise.
<sup>1</sup>Baseline demographic and clinical characteristics were defined at examination 7.
Abbreviations: HDL, high-density lipoprotein; ICV, kg, kilogram; m, meter; NP= neuropsychology; n, number.

### Dietary inflammatory index scores and cognitive decline

Higher (pro-inflammatory) DII scores were linearly associated with accelerated decline in performance on the test of Trail Making B-A after adjustment for age, sex and education lifestyle factors (BMI, total energy intake, and smoking status), cardiometabolic factors (total to HDL cholesterol ratio and the use of anti-cholesterol medication) (Model 2: beta ± standard error = −0.010 ± 0.004, p=0.03) **(Table 2)**. In addition, in comparison to the first quartile (i.e. most anti-inflammatory), we found that the fourth DII score quartile (i.e. pro-inflammatory) was associated with decline in performance on the test of Trail Making B-A (Model 2: quartile 4: −0.033 ± 0.014, p =0.03). And we observed a significant p for trend (Model 2 p=0.02) as well. Further, higher DII scores were linearly related with (less) decline in performance on the test of Similarities after adjustment for Model 1 covariates (Model 1: beta ± standard error = 0.017 ± 0.008, p=0.03) **(Table 2)**. After we additionally adjusted for Model 2 covariates the association did not hold (Model 2: 0.018 ± 0.008, p=0.06). Additionally, in comparison to the first quartile, the third DII score quartile was associated with (less) decline in performance on the test of Similarities (Model 1: quartile 3: 0.095 ± 0.036, p =0.01; Model 2: 0.091 ± 0.038, p= 0.02). And higher DII scores were linearly associated with less decline of global cognition (Model 1: 0.004 ± 0.002, p= 0.02, p for trend= 0.01; Model 2: 0.004 ± 0.002, p= 0.03, p for trend= 0.03) **(Table 2)**. Lastly, in comparison to the first quartile, the fourth DII score quartile was related to (less) decline of global cognition in model 1 (Model 1: quartile 4: 0.018 ± 0.008, p= 0.02; Model 2: quartile 4: 0.018 ± 0.009, p= 0.05). We observed no associations between the DII and annualized change in performance on Logical Memory Delayed Recall, Visual Reproduction Delayed Recall, Trail Making A and the Hooper Visual Organization Test.

**Table 2.** Association between Dietary Inflammatory Index (DII) and Cognitive decline.

| Logical memory delayed, n Correct (n=1608) |  |  |  |  |
| --- | --- | --- | --- | --- |
|  | Model 1 |  | Model 2 |  |
| | $\beta \pm SE^*$ | P-value† | $\beta \pm SE^*$ | P-value† |
| DII Score (cont.) | 0.001 ± 0.009 | 0.93 | 0.006 ± 0.011 | 0.60 |
| **DII Scores (quartiles)** |  |  |  |  |
| Q1 [-4.39, -1.52] (n= 402) | *Reference* |  | *Reference* |  |
| Q2 [-1.52, -0.25] (n= 402) | 0.028 ± 0.042 | 0.50 | 0.042 ± 0.043 | 0.33 |
| Q3 [-0.24, 0.98] (n= 402) | -0.003 ± 0.042 | 0.95 | 0.010 ± 0.045 | 0.82 |
| Q4 [0.98, 3.95] (n= 402) | 0.025 ± 0.043 | 0.56 | 0.051 ± 0.049 | 0.31 |
| *P for trend* |  | 0.74 |  | 0.46 |
| Visual reproduction delayed, n Correct (n=1608) |  |  |  |  |
|  | Model 1 |  | Model 2 |  |
| | $\beta \pm SE^*$ | P-value† | $\beta \pm SE^*$ | P-value† |
| DII Score (cont.) | 0.008 ± 0.007 | 0.26 | 0.006 ± 0.009 | 0.52 |
| **DII Scores (quartiles)** |  |  |  |  |
| Q1 [-4.39, -1.51] (n= 402) | *Reference* |  | *Reference* |  |
| Q2 [-1.49, -0.24] (n= 402) | 0.040 ± 0.034 | 0.23 | 0.041 ± 0.035 | 0.23 |
| Q3 [-0.24, 0.98] (n= 402) | 0.031 ± 0.034 | 0.36 | 0.024 ± 0.036 | 0.50 |
| Q4 [0.99, 3.95] (n= 402) | 0.044 ± 0.034 | 0.20 | 0.031 ± 0.040 | 0.44 |
| *P for trend* |  | 0.26 |  | 0.56 |

Abbreviations: HDL= high-density lipoprotein; NP= neuropsychology; n= number; q= quartile.
Note. Model 1: Age, sex and education.

| Trail Making A, minutes (n=1581) |  |  |  |  |
| --- | --- | --- | --- | --- |
|  | Model 1 |  | Model 2 |  |
| | $\beta \pm SE^*$ | P-value<sup>†</sup> | $\beta \pm SE^*$ | P-value<sup>†</sup> |
| DII Score (cont.) | 0.001 ± 0.001 | 0.35 | 0.001 ± 0.001 | 0.16 |
| **DII Scores (quartiles)** |  |  |  |  |
| Q1 [-4.39, -1.51] (n= 395) | *Reference* |  | *Reference* |  |
| Q2 [-1.51, -0.27] (n= 395) | -0.004 ± 0.003 | 0.14 | -0.004 ± 0.003 | 0.18 |
| Q3 [-0.27, 0.99] (n= 396) | 0.001 ± 0.003 | 0.67 | 0.002 ± 0.003 | 0.46 |
| Q4 [0.99, 3.95] (n= 395) | 0.001 ± 0.003 | 0.74 | 0.003 ± 0.003 | 0.42 |
| P for *trend* |  | 0.36 |  | 0.18 |
| Trail Making B – A, minutes (n=1543) |  |  |  |  |
|  | Model 1 |  | Model 2 |  |
| | $\beta \pm SE^*$ | P-value<sup>†</sup> | $\beta \pm SE^*$ | P-value<sup>†</sup> |
| DII Score (cont.) | -0.005 ± 0.003 | 0.08 | **-0.010 ± 0.004** | **0.03** |
| **DII Scores (quartiles)** |  |  |  |  |
| Q1 [-4.39, -1.52] (n= 386) | *Reference* |  | *Reference* |  |
| Q2 [-1.52, -0.27] (n= 386) | 0.013 ± 0.014 | 0.36 | 0.008 ± 0.014 | 0.56 |
| Q3 [-0.27, 0.97] (n= 386) | -0.003 ± 0.014 | 0.81 | -0.011 ± 0.014 | 0.45 |
| Q4 [0.97, 3.95] (n= 386) | -0.023 ± 0.014 | 0.10 | **-0.033 ± 0.014** | **0.04** |
| P for *trend* |  | 0.06 |  | **0.02** |

Abbreviations: HDL= high-density lipoprotein; NP= neuropsychology; n= number; q= quartile.
Note. Model 1: Age, sex, and education.

| Hooper visual organization, n Correct (n=1562) |  |  |  |  |
| --- | --- | --- | --- | --- |
|  | Model 1 |  | Model 2 |  |
| | $\beta \pm SE^*$ | P-value<sup>†</sup> | $\beta \pm SE^*$ | P-value<sup>†</sup> |
| DII Score (cont.) | 0.004 ± 0.001 | 0.46 | 0.003 ± 0.007 | 0.60 |
| **DII Scores (quartiles)** |  |  |  |  |
| Q1 [-4.39, -1.52] (n= 390) | *Reference* |  | *Reference* |  |
| Q2 [-1.49, -0.27] (n= 391) | 0.026 ± 0.025 | 0.30 | 0.028 ± 0.026 | 0.27 |
| Q3 [-0.25, 0.99] (n= 391) | 0.012 ± 0.026 | 0.64 | 0.010 ± 0.027 | 0.70 |
| Q4 [1.00, 3.95] (n= 390) | 0.035 ± 0.026 | 0.17 | 0.036 ± 0.030 | 0.23 |
| P for *trend* |  | 0.27 |  | 0.36 |
| Similarities, n Correct (n=1614) |  |  |  |  |
|  | Model 1 |  | Model 2 |  |
| | $\beta \pm SE^*$ | P-value<sup>†</sup> | $\beta \pm SE^*$ | P-value<sup>†</sup> |
| DII Score (cont.) | **0.017 ± 0.008** | **0.03** | 0.018 ± 0.009 | 0.06 |
| **DII Scores (quartiles)** |  |  |  |  |
| Q1 [-4.39, -1.52] (n= 403) | *Reference* |  | *Reference* |  |
| Q2 [-1.51, -0.25] (n= 404) | 0.016 ± 0.036 | 0.66 | 0.009 ± 0.036 | 0.81 |
| Q3 [-0.23, 0.98] (n= 404) | **0.095 ± 0.036** | **0.01** | **0.091 ± 0.038** | **0.02** |
| Q4 [0.99, 3.95] (n= 403) | 0.044 ± 0.036 | 0.22 | 0.038 ± 0.042 | 0.36 |
| P for *trend* |  | 0.07 |  | 0.13 |

Abbreviations: HDL= high-density lipoprotein; NP= neuropsychology; n= number; q= quartile.
Note. Model 1: Age, sex, and education.

| Global cognition, weighted score units (n=1527) |  |  |  |  |
| --- | --- | --- | --- | --- |
|  | Model 1 |  | Model 2 |  |
| | $\beta \pm SE^*$ | P-value<sup>†</sup> | $\beta \pm SE^*$ | P-value<sup>†</sup> |
| DII Score (cont.) | **0.004 ± 0.002** | **0.02** | **0.004 ± 0.002** | **0.03** |
| **DII Scores (quartiles)** |  |  |  |  |
| Q1 [-4.39, -1.54] (n= 381) | *Reference* |  | *Reference* |  |
| Q2 [-1.54, -0.28] (n= 382) | 0.003 ± 0.008 | 0.72 | 0.004 ± 0.008 | 0.59 |
| Q3 [-0.28, 0.96] (n= 382) | 0.013 ± 0.008 | 0.09 | 0.014 ± 0.008 | 0.10 |
| Q4 [0.97, 3.95] (n= 382) | **0.018 ± 0.008** | **0.02** | 0.018 ± 0.009 | 0.05 |
| P for trend | **0.01** |  | **0.03** |  |

Abbreviations: HDL= high-density lipoprotein; NP= neuropsychology; n= number; q= quartile.
Note. Model 1: Age, sex, and education.
Model 2: Model 1 + Time from Exam 7 to NP Evaluation, body mass index, smoking status, total energy intake, total cholesterol to HDL cholesterol ratio and the use of anti-cholesterol medication. \*The beta estimates give the standard deviation unit difference in each respective outcome per one unit increase in the DII score. <sup>†</sup>A P-value <.05 was considered statistically significant.

We observed significant interactions between the DII (continuous and categorically) and sex in their associations with Hooper Visual Organization Test performance **(eTable 3 in the supplement)**. Following stratification of results, higher DII scores were linearly associated with (less) decline in performance on the Hooper visual organization Test among men (Model 2: - 0.022 ± 0.010,p=0.02, p=0.02, p for trend =0.01), but not among women (Model 2: −0.011 ± 0.009, p=0.23) **(eTable 3 in the supplement)**.

## Discussion

In the present study we evaluated whether higher DII scores were associated with cognitive decline in the large community-based FHS Offspring cohort. We found associations between higher pro-inflammatory DII scores and (less) cognitive decline among men and women combined. And we reported that the relationships between higher DII scores and executive function and processing speed may be modified by sex.

We take our findings with caution as we would expect higher DII scores to be associated with accelerated cognitive decline. We recently published that higher DII scores are associated with increased risk of all-cause and AD dementia in the same study population (Hazard ratio (HR) 1.21, 95% confidence interval (CI) 1.10 to 1.33, p<0.001; HR 1.20, 95% CI 1.07 to 1.34, p=0.001, respectively)^11^. In addition, we reported that higher DII scores relate to smaller total brain volume (beta ± standard error: −0.16 ± 0.03; P < .0001), smaller total gray matter volume (- 0.08 ± 0.03; p= 0.003) and larger lateral ventricular volume (0.04 ± 0.02; p = 0.03). These relationships are all as expected. We could speculate that our study sample has increased cognitive reserve (∼72% had high-school degree or higher) and, therefore, they might have a slow decline in cognitive test performance over time despite accumulating neuropathology.

### Findings in other studies

*Global cognition* – We observed that higher DII scores were associated with less decline in global cognition over a maximum of 11 years of follow-up. The supplementation with antioxidant vitamins and minerals (SU.VI.MAX) cohort investigated DII in relation to a composite global cognition score 13 years apart, showed a relationship between pro-inflammatory DII scores and worse global cognition^18^. In addition, multiple cross-sectional studies reported relationships between higher DII scores and worse global cognition^14–17^. In contrast, in the Nurses’ Health Study (NHS) (n=16,056) we investigated relationships between an Empirical Dietary Inflammatory Pattern (EDIP) and a composite global cognition score, and the Telephone Interview for Cognitive Status (TICS) over six years of follow-up. After adjustment for demographic, lifestyle, and disease (related) risk factors we observed no associations between EDIP and cognitive aging^22^. We speculated that our null findings may be due to the high educational level of the nurses and moderate follow-up time (6 y) of the cohort. Further, two cross-sectional studies also observed no significant relationships between (energy-adjusted) DII and composite global cognition scores^19^ ^23^.

*Executive function* – In contrast to our findings SU.VI.MAX observed no association between DII and executive function^18^.

*Memory* – We observed no relationship between DII and memory in the present study and the NHS study^22^. This is in line with our previous reported null finding between DII and hippocampal volume in the same study population ^13^. The SU.VI.MAX, however, reported a significant association between higher DII scores and worse verbal memory, but only among individuals within the two highest pro-inflammatory quartiles as compared to the lowest pro-inflammatory quartile^18^. In addition, two cross-sectional studies observed relationships between higher DII and worse episodic memory, semantic-based memory, and executive function and working-memory combined (i.e. Digit Symbol Substitution Test)^20^ ^21^. However, residual confounding may be present as no adjustments were made for cholesterol lowering medication and cholesterol blood concentrations, which may (partly) explain the relationship.

*Visuospatial* – In addition to global cognition, we take our finding that higher DII scores were associated with less decline in visuospatial domain with caution. The Washington Heights, Hamilton Heights, and Inwood Columbia Aging Project (WHICAP) reported that higher a priori–defined inflammation-related nutrient pattern (INP) scores were associated with worse visuospatial test performance^23^.

*Verbal comprehension* – To the best of our knowledge no study has yet evaluated DII with this cognitive domain.

To date, most observational studies that investigated DII in relation to cognitive aging have been carried out in cross-sectional studies. We encourage cohorts that have repeated measures of robust neuropsychological tests over a long follow-up time to replicate our findings. Thus, research is needed to elucidate whether inflammatory content of diet affects age related cognitive decline.

### Mechanisms

Systemic inflammation is one of the hypothesized contributary pathways leading to cognitive aging and brain disease.^32^ ^33^ As mentioned earlier, a driving factor of systemic inflammation is ‘inflammaging’, which activates microglial macrophages to produce pro-inflammatory cytokines (e.g. CRP, interleukin-1β, IL-6, and TNF- α.^11^ ^33^ Production of these cytokines can cause neuronal death, cerebral small vessel disease, neurodegeneration, and brain atrophy.^11^ ^33^ Indeed, a previous study performed by members of our team showed a relationship between higher concentrations of inflammatory marker plasma soluble CD14 (sCD14) – a suggested microglial inflammatory response regulator - and worse global cognition and worse performance on the test of Similarities (cross-sectionally), accelerated cognitive decline (i.e. executive function), larger total brain volume atrophy and an increased risk for all-cause dementia.^34^ In addition, our team reported associations between higher levels of cytokines and larger white matter hyperintensity volumes, more cerebral microbleeds and silent cerebral infarcts, all markers of cerebral small vessel disease.^35^ Further, with regard to reduced brain volume caused by cytokines, our group showed that higher levels of cytokines, IL-6, osteoprotegerin and TNF- α, were related with smaller total brain volume.^36^

#### Strengths and limitations

Strengths of this study include our large population-based sample, a maximum follow-up time of 11 years, a validated FFQ, the creation of an averaged DII over a maximum of three time points to estimate DII over a maximum of 10 years follow-up and the ability to comprehensively investigate cognitive aging by using a wide range of cognitive tests. Additionally, we were able to adjust for many important demographic, lifestyle, and disease risk factors for cognitive aging.

However, we acknowledge that the present study has limitations. First, to assess DII scores based on dietary intake we used a FFQ, which is subject to measurement error and recall bias. To mitigate the effects of recall bias, we adjusted for total energy intake in model 2 to account for potential systematic measurement error.^37^ We acknowledge the possible presence of non-differential misclassification while using this dietary assessment method, which may have led to bias towards the null.^38^ Second, in the FFQ 36 out of 45 DII components were available. However, our quantity of components is at the upper limit (DII component range across studies: 23-45) of studies that have investigated the DII and cognitive aging.^14–21^ Third, the present study is observational, which precludes conclusions about the causality of the observed associations due to potential reverse causality. However, the DII score was constructed over three consecutive visits over a maximum of 10 years of follow up, which makes such a reverse causation bias less likely. Lastly, the generalizability of our findings to other races/ethnicities may be limited as individuals included in our study were mainly white individuals of European ancestry. However, the DII accounts for that to a certain extent by including a mean and standard deviation of a representative world database, which we used in our calculation.^10^

## Conclusion

In conclusion, in our community-based sample, higher DII scores were associated with (less) cognitive decline. We take our findings with caution as we previously have shown a relationship between higher DII scores and risk of incident dementia. Further, most studies that found a relationship between DII and cognition were cross-sectional and have used less sensitive testing. Future longitudinal studies with sensitive neuropsychological test measures are encouraged to elucidate whether a longitudinal relationship between higher DII scores and age-related cognitive decline exists.

## Statements

### Data access

All authors confirm to have full access to all the data in the study and accept responsibility to submit for publication.

### Data sharing

Data are available on request for bone fide investigators from managing institution of the Framingham Heart Study https://www.framinghamheartstudy.org/fhs-for-researchers/.

## Data Availability

Data will be available for researchers with an approved FHS research proposal and IRB approval and who have signed a data sharing agreement. Outside investigators with approved research proposals and Institutional Review Board protocols can access genomic and phenomic data via multiple mechanisms as outlined below.
Beginning in 2007, extensive phenotype genotype data sets for all data in the SHAReproject has been available to investigators who submit
a) a short research proposal
b) evidence of a local IRB approved project
c) an executed DMDA data materials and distribution agreement
d) a computer security plan. The DMDA outlines issues about participant confidentiality, computer security and intellectual property. The proposals will be reviewed by NHLBI staff with a very short turnaround time. The protocols and data are updated frequently.
Privacy, rights, and confidentiality of human research participants will be protected by de-identifying the data and ensuring that no personal health information can be linked to a research participant.

https://www.framinghamheartstudy.org/

## Acknowledgement

We thank the FHS participants for donating their time to our research.

## Conflicts of interest disclosures

Dr. Melo van Lent is chair of the Alzheimer’s Association ISTAART Nutrition Metabolism and Dementia Professional Interest Area; Dr. Paul Jacques is part of the Danone North America Essential Dairy and Plant-Based Advisory Board; Hannah Gokingco declarations of interest: none; Dr. Claudia Satizabal declarations of interest: none; Dr. Sokratis M. Charisis declarations of interest: none; Dr. Changzheng Yuan declarations of interest: none; Dr. Ramachandran S. Vasan declarations of interest: none; Dr. Sudha Seshadri declarations of interest: none; Dr. Alexa Beiser declarations of interest: none; Dr. Jayandra J. Himali declarations of interest: none; and Dr. Mini E. Jacob declarations of interest: none.

## Funding sources

The Framingham Heart Study was supported by the National Heart, Lung, and Blood Institute (contract no. N01- HC-25195, HHSN268201500001I and no. 75N92019D00031); and the National Institute on Aging (NIA) (R01 AG054076, R01 AG049607, R01 AG033193, U01 AG049505, U01 AG052409, U01 AG058589, RF1 AG059421)); and by grants from the National Institute of Neurological Disorders and Stroke (NS017950 and UH2 NS100605). Dr. Melo van Lent received funding provided by the NIH-NIA (R03 AG067062-01 and R03 AG087437-01) and Alzheimer’s Association Research Grant (AARG-1241954) and Alzheimer’s Association Research Fellowship grant (AARF-22-918316) to support this research project. In addition she is supported by NIH-NIA grants 1RF1AG059421 and 1P30 AG066546-01A1; Funds from the USDA Agricultural Research Service Agreement (No. 58-8050-9-004) supported in part the collection of dietary data for this project and the efforts of Dr. Paul Jacques. In addition, Dr. Paul Jacques is supported by R01 AG059011-01A1, R01 DK134533-01A1 and the Institute for the Advancement of Food and Nutrition Sciences; Dr. Satizabal was supported by a New Investigator Research Grant to promote Diversity from the Alzheimer’s Association (AARGD-16-443384) and also receives support from NIA (R01 AG059727 and R01 AG082360); Dr. Changzheng Yuan is supported by the Alzheimer’s Association (AARG-22- 928604) and the University Global Partnership fund; Drs. Claudia L. Satizabel, Sudha Seshadri and Jayandra J. Himali are partially supported by the South Texas Alzheimer’s Disease Center (1P30AG066546-01A1) and The Bill and Rebecca Reed Endowment for Precision Therapies and Palliative Care; Dr. Seshadri is also supported by 1RF1AG059421 and by an endowment from the Barker Foundation as the Robert R Barker Distinguished University Professor of Neurology, Psychiatry and Cellular and Integrative Physiology; Dr. Himali is also supported by AG062531 and by an endowment from the William Castella family as William Castella Distinguished University Chair for Alzheimer’s Disease Research. The funding agencies had no role in study design; in the collection, analysis and interpretation of data; in the writing of the report; and in the decision to submit the article for publication.

## Notes

### Competing Interest Statement

Dr. Melo van Lent is chair of the Alzheimers Association ISTAART Nutrition Metabolism and Dementia Professional Interest Area. Dr. Paul Jacques is part of the Danone North America Essential Dairy and Plant-Based Advisory Board. Hannah Gokingco declarations of interest: none. Dr. Claudia Satizabal declarations of interest: none. Dr. Sokratis M. Charisis declarations of interest: none. Dr. Changzheng Yuan declarations of interest: none. Dr. Ramachandran S. Vasan declarations of interest: none. Dr. Sudha Seshadri declarations of interest: none. Dr. Alexa Beiser declarations of interest: none. Dr. Jayandra J. Himali declarations of interest: none. and Dr. Mini E. Jacob declarations of interest: none.

### Author Declarations

The institutional review board at Boston University Medical Center gave ethical approval for this work

