## Supplementary material for "Dietary inflammatory index scores and cognitive aging: results from the Framingham heart Study offspring cohort"

### Supplementary eTable 1. Description of Dietary Inflammatory Index calculation

| **Description** |
| --- |
| The DII is based on a set of literature-based dietary-component- specific inflammatory effect scores, dietary intake data from the population under study, and a representative world database that provides a mean and standard deviation for consumption of each dietary component in the global population.[1] These then become the multipliers to express an individual's exposure relative to the 'standard global mean' as a Z-score: subtracting the 'standard global mean' (i.e. from the world database) from the amount reported by the study participants and dividing this value by the world standard deviation. To minimize the effect of 'right skewing', this value is converted to a percentile score. To achieve a symmetrical distribution with values centered on 0 (null) and bounded between −1 (maximally anti- inflammatory) and +1 (maximally pro-inflammatory), each percentile score is doubled and then '1' is subtracted. The centered percentile value for each dietary component is then multiplied by its respective 'overall dietary component-specific inflammatory effect score' to obtain the 'dietary component specific DII score'. Finally, all of the 'dietary component-specific DII scores' are summed to create the 'overall DII score' for an individual, higher scores indicating pro- inflammatory DII scores).[1] |

NOTE. References

1. Shivappa N, Steck SE, Hurley TG, et al. Designing and developing a literature-derived, population-based dietary inflammatory index. Public Health Nutr 2014;17(8):1689-96.

### Supplementary eTable 2. Description of covariates

| Covariate | Description |
|---|---|
| Body mass index | BMI was defined as weight (in kilograms) divided by the square of height (in meters). |
| Cholesterol | Fasting blood samples at examination cycle 7 were drawn from participants after an overnight fast. Total cholesterol and high-density lipoprotein (HDL) cholesterol concentrations were measured directly using standardized assays. |
| Education | Education was categorized into 3 groups (up to completion of 12 years of education leading to high school degree but no college; some college; college degree). |
| Anti-cholesterol medication | Lipid lowering medication (i.e. statins, fibrates, resins, niacin or nicotinic acid, and other anti-cholesterol drugs) usage was categorized into users and non-users. |
| Physical activity | Physical activity was self-reported using the physical activity index [3]. |
| Smoking status | Indicator variables were used for current smoking. |
| Total energy intake | Total energy intake was estimated from the Food Frequency questionnaire. |

NOTE. References

1. Elosua R, Ordovas JM, Cupples LA, et al. Association of APOE genotype with carotid atherosclerosis in men and women: the Framingham Heart Study. J Lipid Res 2004; 45(10): 1868-75.

2. Hixson JE, Vernier DT. Restriction isotyping of human apolipoprotein E by gene amplification and cleavage with HhaI. J Lipid Res 1990; 31(3): 545-8.

3. Kannel WB, Sorlie P. Some health benefits of physical activity. The Framingham Study. Arch Intern Med 1979; 139(8): 857-61.

eTable 3. Interaction in the association between Dietary Inflammatory Index (DII) and Cognitive tests (results are *P*-values)

| | **Continuous DII Score** | **Categorical DII Score** |
|---|---|---|
| | **Interaction Variables*** | |
| | **Sex** | **Sex** |
| Logical memory delayed, n Correct | 0.27 | 0.38 |
| Visual reproduction delayed, n Correct | 0.63 | 0.71 |
| Trail Making A, min | 0.32 | 0.30 |
| Trail Making B-A, min | 0.50 | 0.49 |
| Hooper visual organization, n Correct | **0.06** | **0.097** |
| Similarities, n Correct | 0.52 | 0.30 |
| Global cognition, weighted score units | 0.77 | 0.54 |

Abbreviations: HDL= high-density lipoprotein; min= minutes.

Note. Model 2: Age, sex, education, body mass index, smoking status, total energy intake, total cholesterol to HDL cholesterol ratio and the use of anti-cholesterol medication. . *A P-value <.10 was considered statistically significant..

eTable 4. Association between continuous Dietary Inflammatory Index (DII) and Hooper visual organization, stratified by sex.

| DII Score | Men (n =696) | | | Women (n =866) | |
|---|---|---|---|---|---|
| | Hooper visual organization, n Correct | | | Hooper visual organization, n Correct | |
| | β ± SE | P-value‡ | | β ± SE | P-value‡ |
| DII Score (cont.) | **0.022 ± 0.010** | **0.02** | | -0.011 ± 0.009 | 0.23 |
| **DII Score in quartiles** | | | **DII Score in quartiles** | | |
| Q1 (n= 165) | *Reference* | | Q1 (n= 225) | *Reference* | |
| Q2 (n= 184) | -0.004 ± 0.038 | 0.92 | Q2 (n= 207) | 0.056 ± 0.036 | 0.10 |
| Q3 (n= 177) | 0.067 ± 0.040 | 0.09 | Q3 (n= 214) | -0.035 ± 0.038 | 0.35 |
| Q4 (n= 170) | **0.084 ± 0.043** | **0.05** | Q4 (n= 220) | 0.002 ± 0.041 | 0.97 |
| P for trend | **0.02** | | P for trend | 0.50 | |

Abbreviations: HDL= high-density lipoprotein; n= number.

Note. Model 2: Age, education, body mass index, smoking status, total energy intake, total cholesterol to HDL cholesterol ratio and the use of anti-cholesterol medication. †The beta estimates give the standard deviation unit difference in each respective outcome per one unit increase in the DII score. ‡A P-value <.05 was considered statistically significant.
